# Experiences and observations from a care point for displaced Ukrainians: a community case study in Antwerp, Belgium

**DOI:** 10.1101/2024.01.17.24301399

**Authors:** Stefan Morreel, Veronique Verhoeven, Hilde Bastians, Katrien Monten, Josefien van Olmen

## Abstract

**Background:** 7307 Ukrainians refugees moved to Antwerp, Belgium during the study period (01/04/2023-31/12/2023). The city’s administration set up three care centers where these people were introduced in the Belgian primary care system, a medical file was created, and acute/preventive/chronic care was delivered. This community case study analyses the organization and contents of care and reflects upon its meaning for the mainstream health care system.

**Methods:** This is an observational study using routine electronic medical record data to measure uptake of care. For a sample of 200 subjects, a retrospective chart review was conducted in order to extract data from patient records.

**Participants:** all refugees with a medical file at one of the three participating care centers.

**Main Outcomes:** For the observational study, 2261 patients were reached (30% of the potential users) and 6450 contacts were studied. The nurses (including midwives) did 4929/6450 (76%) of all consultations, the GPs 1521/6450 (24%). Out of the 4929 nurse consultations, 955 (19%) were followed by another nurse consultation and 866 (18%) by a GP consultation. In the structured case reviews most contacts concerned acute problems (609/1074, 57%). The most prevalent reasons for encounter and diagnoses concerned typical primary care issues. The nurses were able to manage half of the cases independently (327, 55%), referred to the GP in 37% (N=217), and consulted a GP (live, by telephone or a dedicated app) for 8% (48). GPs mostly prescribed drugs, referred to a medical specialist and advised over-the-counter drugs while nurses more often advised over-the-counter drugs (mostly paracetamol, nose sprays and anti-inflammatory drugs), gave non-medical advice or ordered lab tests.

**Discussion:** The medical care points delivered mostly typical acute primary care in this first phase, with a key role for nurses. These results will inform policy makers on the use of primary care centers for newly arriving patients in times of crisis. A nurse first model seems feasible and efficient, but evaluation of safety and quality of care are needed. Once the acute phase of this crises fades away, questions about the comprehensiveness, continuity, and integration of care for migrants remain relevant.

## 1 Introduction

International crises, such as war, persecution, and natural disasters force people to move from their home environment. Other countries need to prepare themselves to welcome refugees and provide shelter. This means that destination countries need to think about how to address needs of newcomers for health and wellbeing which includes housing, education, employment, health, social inclusion and care, and other social services.(1) The influx of forced migrants tends to occur in waves that are partially unpredictable. Since destination countries tend to scale down their refugee support in times of low arrivals, the rise is – every time again - perceived as an acute crisis for which administrative and support systems are insufficiently prepared.(2) The response of government actors therefore bears an element of pressure which requires an acute investment of time and money. At the same time, it can unleash new energy and out of the box solutions. In this way, refugee crises resemble other crises that put stress on healthcare systems such as COVID-19.

The organization of health care for forced migrants in Europe has gained increasing attention in recent decades, partly because of the human catastrophes during and after their journey depicted in the media [add ref] and partly because of the framing of migrant health as a health security issue with impact on the general population).(3) Barriers in access to health care are highly prevalent in migrant populations, and can lead to underusage and misuse of health care with potentially increased healthcare costs. The barriers in access and quality of care at the operational level have been well described from both user (4) and provider (5) perspective. These include: language and communication barriers, differences in sociocultural norms and illness perceptions, low health literacy and the failure to understand or navigate in new health care systems, perceived discrimination and low trust in the system, and precarious circumstances interfering with health and health care access such as social deprivation, trauma, and lack of insurance coverage.(6) Despite these studies identifying clear gaps, there is little knowledge on system-level interventions to improve the performance of health care services that would benefit both forced migrants and host societies.(7) For instance how to organize care that is accessible and addresses actual needs of forced migrants, which data to collect to optimize patient care, who is best placed to provide care?

The aim of this community case study is to analyze a local health initiative set up to address the needs of a sudden influx of forced migrants (from Ukraine). It addresses several subjects from a health service perspective: Organization of care delivery; Process outcomes in terms of delivery of care and population reached; Content of care. The results of this study are relevant for the future care of the target population and might inspire health care organization for other target populations and crises context.

The studied local health initiative originates from a government initiative to tackle the medical needs of an (at that time) expected large influx of Ukrainian refugees. The Belgian federal government (responsible for most of the crisis management and the healthcare system) asked the Flemish regional government (responsible for prevention and the organization of primary care) to set up local initiatives to address health needs of these refugees. These local initiatives consisted of care points (“Zorgpunten” in Dutch) which had to be set up by local primary care zones. These zones are a collaboration of the local government, healthcare workers and social welfare organizations with the aim to facilitate healthcare professionals to deliver person centered, accessible and high-quality primary care.(8) In the city of Antwerp (part of Flanders), four primary care zones started a combined initiative which will further be called the “care points”. These Antwerp care points emerged from a similar local initiative to combat COVID-19.(9) By coincidence, the end of the COVID-19 crisis coincided with the arrival of the first Ukrainian refugees. The primary care zones had an exceptional liberty in organizing these care points. They were financed per capita in contrast to most of the Belgian Healthcare system which has a fee for service payments.

In this paper we describe the initiative and report the results of a retrospective observational study using routine data from the electronic files to assess the process and content of care. For a sample of 200 subjects, an additional retrospective chart review was conducted to complement and deepen this data.(10)

For the observational study, all recorded contacts at the care points during the study period (31/03/2022-31/12/2024) were assessed for eligibility (N = 8346). Contacts for non-Ukrainian citizens, no-shows, questions answered by the administrative staff, telephone consultations and consultations beyond the study period were excluded. Data were extracted from the care point’s electronic patient files (Mediris Multi®) by a staff member of the care points. Patient names, healthcare professional names and healthcare professional registration numbers were pseudonymised using the irreversible SHA 256 algorithm by the same staff member. Patient’s date of birth was replaced by age after data cleaning. For most patients, a structured assessment of their prevention needs primarily focused on tuberculosis screening and vaccinations was conducted, mostly during their initial encounter (intake). The authors also intended to study this preventive assessment, but these data were unsuitable for analysis due to registration flaws. While these contacts were included in the overall number of consultations, specific prevention outcomes were not analyzed.

For the retrospective chart review, a care point administrative staff member extracted a random sample of 200 patient files. This staff member replaced the names of the professionals by their profession (MD, nurse, midwife, or administrative staff), erased the name of the patient and replaced the birthdate by the birthyear. Author SM coded the first 10 files, after which all authors except KM drafted a code book. This code book was evaluated on ten more files jointly coded by the same authors. This code book (in Dutch) is available upon request. A single author coded each file. Although most consultations involved one problem (one subcontact), some consultations consisted of several subcontacts concerning distinct clinical problems (e.g., a patient came to discuss arterial hypertension and a rash). Data fields included timestamp of the consultation, discipline of caregiver, type of contact (new problem, follow-up, intake, prevention only and technical procedure only), chronicity (chronic or acute), reason for encounter (code according to the International Catalogue of Primary Care version 2, ICPC-2), diagnosis (ICPC-2) and actions (see Table 4.). The reason for encounter and the diagnosis were not restricted to strictly medical subjects, procedures or administrative tasks could also be registered. In case of a nurse consultation, the variable interprofessional workflow (nurse works independent, nurse refers to MD or nurse asks MD advice) was assessed as well. The preventive assessment contacts were excluded from the retrospective chart review unless they also comprised non-preventive data.

This study was reviewed and approved by the Institutional Review Board of the Antwerp University hospital (project ID 3952) on 14/11/2022.

Raw data for the retrospective chart review was collected using a structured spreadsheet in Microsoft Excel®. All data was analyzed using JMP Pro ® version 17. Only descriptive statistics were used.

## 2 Context

### 2.1 The Ukrainian refugee crisis

From the start of the war between Russia and Ukraine in February 2021 till the end of 2023, 7.9 million refugees had fled Ukraine.(11) Most of them fled to neighboring counties such as Poland. Unlike most other refugees, Ukrainians are treated as EU-citizens in the entire EU giving them the right of temporary protection. These rights include social rights and access to the health insurance.(12, 13) EU health care systems are trying to adjust to the influx of people from Ukraine and the related health care needs, such as physical and mental trauma related to the war .(14) Data about the health and health care provided to Ukrainian refugees are so far scarce, hampering response and future planning.(15)

### 2.2 Community Context

In Belgium, 65 000 Ukrainians refugees arrived by the end of 2023. The majority (62%) was female, 34% were minors. Only 16% demanded shelter, the other 84% found a shelter themselves (mostly friends or relatives).(11) Ukrainian refugees needed to register themselves to obtain social rights including health insurance. The Belgian health system covers almost the entire population for a wide range of health services. Residents must be affiliated to a sickness fund; and contributions are proportional to income. Belgium is among the top ten spenders on health across EU countries, up to 10.7% of its GDP in 2019.(16) The increasing number of people with chronic diseases and staff shortages lead to overburdening of the primary care system in Belgium especially in larger cities, resulting in patient stops and waitinglists.(17)

The current community case is situated in Antwerp, a city with a majority of its 532 000 people having roots outside of Belgium .(18) In 2022, 7307 Ukrainians registered themselves at the municipality (59% female, average age 29), although the actual figure is probably higher as some might have skipped registration. No reliable data on the departing number of Ukrainians (either travelling further or returning to their homes) was available.

## 3 Key programmatic elements

### 3.1 Organization of care

The routine primary care system already being overstretched, the city of Antwerp decided to organize medical care for this large new population in a separate system. Three care points with complementary functions were set up spread out over the city. A first care point was situated in the acute refugee shelter set up in March 2022 to organize the first welcome and a medical/social intake of new arrivals. This center was closed on 1/11/2022 because the influx became more gradually and predictable. Most of the refugees found a home within the community, either with or without support from the city’s administration. For them, a second ambulatory care point was set up on 31/03/2023. The large influx of people pushed the city to create additional accommodation facilities, in the form of an emergency container village at the edge of the city for approximately 700 people. This location hosted the third care point set up on the same date.

The care points were staffed by nurses hired by the city of Antwerp and by independent GPs who worked a limited number of hours per week in addition to their routine work. GPs were remunerated on an hourly basis. The person in charge of the care points was a senior nurse with experience in refugee care. Most of the nurses hired also had experience of working in refugee settings. Apart from providing care, they were actively involved in on-the-job peer training of other staff in the care points. These care points were free of charge to the patients. Nurses were the first point of contact. Patients could access a GP only after a nurse had done the first intake and decided this was necessary. Some chronic patients were actively transferred to regular primary or secondary care, but no data is available on these transfers.

The city aimed for optimal access to both the preventive and curative care serviced provided by the care points. At the moment of registration at the city’s admin and social services, people were asked to sign an informed consent. In this informed consent they gave permission to be contacted for preventive care and a medical intake. Complementarily, the city distributed information on the care points through leaflets, text messages, a Dutch website and through explanation during civic integration courses. Care points would then contact people to offer an intake that offered preventive screening and care and curative care if necessary. Primary care professionals were also informed so that they could refer people.

### 3.2 Process outcomes and content of care: observational study

By the end of this study, 6722 out of 7307 (91%) of the registered refugees signed the informed consent, only two explicitly refused. The remaining 583 did not sign, most likely because the administration forgot to ask them, or the signature got lost.

In the period 31/03/2023 to 31/12/2022, 2261 patients were registered at the Care Points (30% of all registered refugees, 34% of those who signed the informed consent). The total number of health care contacts registered in that period was 8346. Contacts for non-Ukrainian citizens (N=24), no shows (N=117), questions answered by the administrative staff (1152), telephone consultations (N=2) and consultations beyond the study period (N=558) were excluded leading to a total of 6450 studied contacts. The mean number of contacts was 2,87 (Standard Deviation 2.71, median 2). Mean age of the was 27 years; 59% were female. The nurses did 4863/6450 (75%) of all consultations, the GPs 1521/6450 24%, and midwives 66/6450 (1%). Because of the small number of midwife consultations, consultations by midwifes and nurses were taken together in the analyses. Reporting them separately could compromise the anonymity of this report.

Thirty-two GPs performed between 2 and 191 consultations (mean 48, standard deviation 57). Six GPs did more than 100 consultations each. A total of 38 nurses executed between 1 and 806 consultations (mean 130, standard deviation 197), 12 nurses did more than 100 consultations each.

To analyze the interprofessional patient flows, we studied those consultations where the same patient was seen twice within seven days. Out of the 4929 nurse consultations, 955 (19%) were followed by another nurse consultation and 866 (18%) by a GP consultation. Out of the 1521 GP consultations, 281 (18%) were followed by a nurse consultation and 136 (9%) by another GP consultation.

### 3.3 Process outcomes and content of care: retrospective chart review

A total of 991 consultations were coded concerning 193 different patients. Seven of the 200 randomly selected patient files were either empty or duplicates. The subpopulation of 193 patients was demographically similar to the overall population of 2261 patients with a mean age of 28 years and 56% females. In 69 consultations, people mentioned numerous unrelated complaints which were coded as subcontacts leading to a total of 1073 contacts. Each author except KM coded at least 200 contacts. The majority of the contacts (624/992 or 63%) were conducted by nurses, 356/992 (36%) by MDs and 3/992 (0%, further analyzed in the group of nurses) by midwives. For nine contacts, the caregiver was unknown. The proportion of nurse contacts (63%) was lower in the retrospective chart review as compared to the observational study population (75%), most likely because preventive consultations (which were all done by the nurses) were excluded from this analysis.

Most consultations concerned acute problems (609/1074, 57%). Half of the contacts (546, 51%) concerned a new complaint, 388 (36%) were follow up, the remaining 139 (13%) concerned an intake consultation, non-routine prevention actions and technical procedures such as injections. See Table 1 for an overview of the most common reasons for encounter. Most contacts concerned respiratory complaints (193, 18% with coughing as the most frequent complaint), procedures and administration (165, 15% with request for prescriptions as the most frequent subject), general and unspecified complaints including (134, 12% with fever as the most frequent subject), and musculoskeletal complaints (108, 10% with backpain as the most frequent subject).

**Table 1.** Reasons for encounter at the care points split into ICPC-2 chapters. Source: structured case review.

| Reason for encounter (ICPC-2 Chapter) | N | Column % |
| --- | --- | --- |
| Respiratory | 193 | 18% |
| Procedures and Administration | 165 | 15% |
| General and Unspecified | 134 | 12% |
| Musculoskeletal | 108 | 10% |
| Skin | 77 | 7% |
| Digestive | 59 | 5% |
| Cardiovascular | 54 | 5% |
| Missing | 49 | 5% |
| Female Genital | 41 | 4% |
| Neurological | 35 | 3% |
| Ear | 32 | 3% |
| Endocrine/Metabolic and Nutritional | 31 | 3% |
| Psychological | 29 | 3% |
| Urological | 21 | 2% |
| Eye | 18 | 2% |
| Pregnancy, Childbearing, Family Planning | 11 | 1% |
| Male Genital | 8 | 1% |
| Blood, Blood Forming Organs, and Immune Mechanism | 6 | 1% |
| Social Problems | 2 | 0% |
| All | 1073 | 100% |

**ICPC-2: International Catalogue of Primary Care version 2**

We compared the final coding of evaluation and of actions of GPs and nurses which illustrate the difference in type of actions of both professions. See Table 2 for an overview of the GP’s most common diagnosis which were logically similar to the reasons for encounter, the same applies to the nurse’s diagnoses (see Table 3). In total, 819 actions were coded; multiple actions were possible for a single subcontact. The management of the cases was different for nurses and GPs. GPs mostly prescribed drugs, referred to a medical specialist and advised over-the-counter drugs while nurses more often advised over-the-counter drugs (mostly paracetamol, nose sprays and anti-inflammatory drugs), gave non-medical advice (e.g. “Rinse your nose and take a spoon of honey”) or ordered lab tests (mostly point of care COVID tests). For 385/1073 contacts (36%), no information on actions was recorded. See tables 4 and 5 for more details.

**Table 2.**
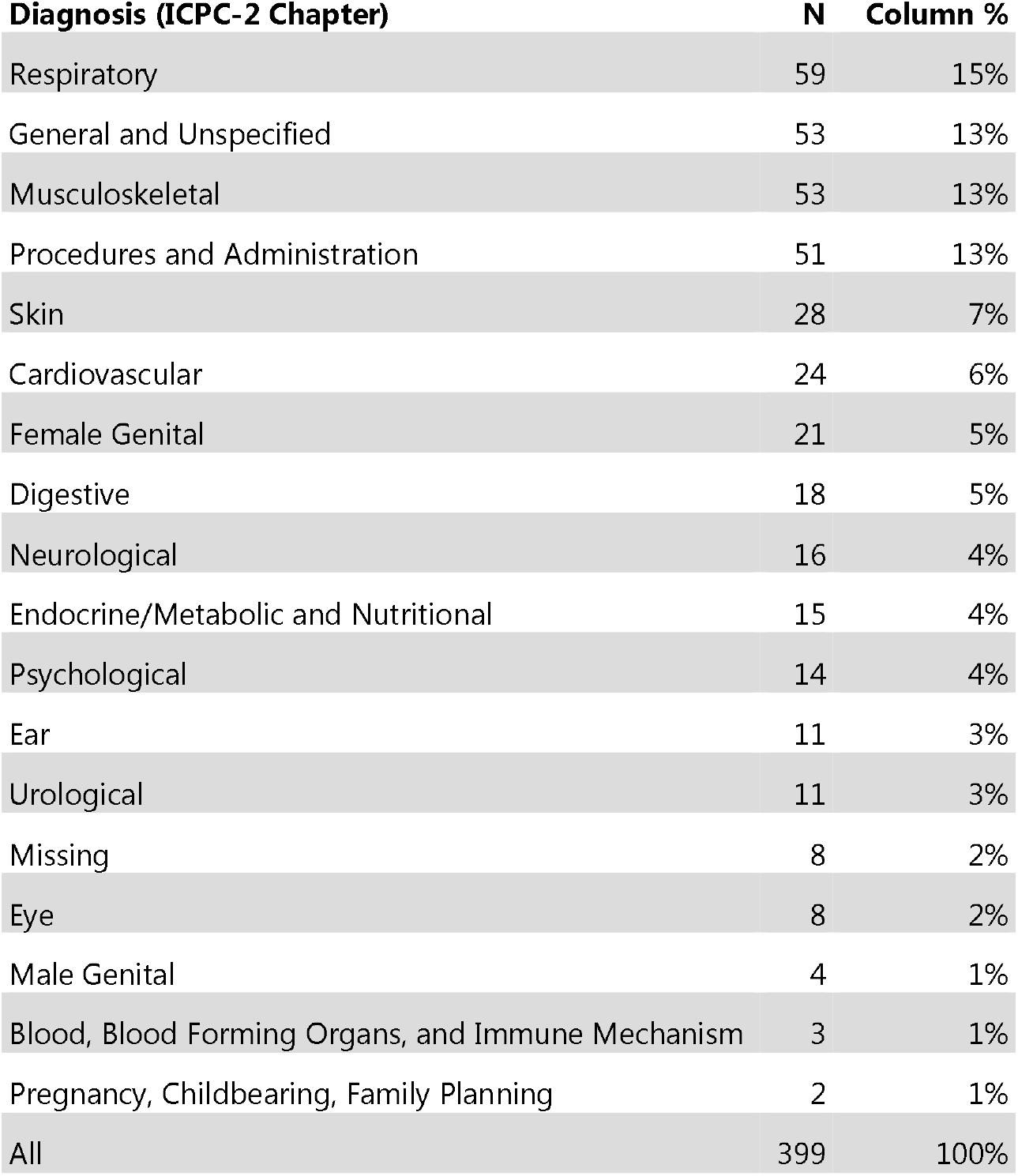
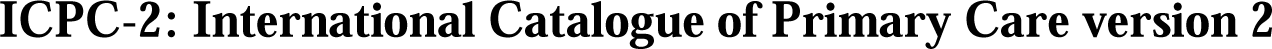
GP diagnoses at the care points split into ICPC-2 chapters. Source: structured case review.

| Diagnosis (ICPC-2 Chapter) | N | Column % |
| --- | --- | --- |
| Respiratory | 59 | 15% |
| General and Unspecified | 53 | 13% |
| Musculoskeletal | 53 | 13% |
| Procedures and Administration | 51 | 13% |
| Skin | 28 | 7% |
| Cardiovascular | 24 | 6% |
| Female Genital | 21 | 5% |
| Digestive | 18 | 5% |
| Neurological | 16 | 4% |
| Endocrine/Metabolic and Nutritional | 15 | 4% |
| Psychological | 14 | 4% |
| Ear | 11 | 3% |
| Urological | 11 | 3% |
| Missing | 8 | 2% |
| Eye | 8 | 2% |
| Male Genital | 4 | 1% |
| Blood, Blood Forming Organs, and Immune Mechanism | 3 | 1% |
| Pregnancy, Childbearing, Family Planning | 2 | 1% |
| All | 399 | 100% |

**ICPC-2: International Catalogue of Primary Care version 2**

**Table 3.** Nurse diagnoses at the care points split into ICPC-2 chapters. Source: structured case review.

| Diagnosis (ICPC-2 Chapter) | N | Column % |
| --- | --- | --- |
| Respiratory | 134 | 20% |
| Procedures and Administration | 113 | 17% |
| General and Unspecified | 80 | 12% |
| Musculoskeletal | 55 | 8% |
| Skin | 48 | 7% |
| Missing | 41 | 6% |
| Digestive | 41 | 6% |
| Cardiovascular | 30 | 5% |
| Ear | 21 | 3% |
| Neurological | 19 | 3% |
| Female Genital | 18 | 3% |
| Endocrine/Metabolic and Nutritional | 15 | 2% |
| Psychological | 14 | 2% |
| Eye | 10 | 2% |
| Urological | 9 | 1% |
| Pregnancy, Childbearing, Family Planning | 8 | 1% |
| Male Genital | 4 | 1% |
| Blood, Blood Forming Organs, and Immune Mechanism | 3 | 0% |
| Social Problems | 2 | 0% |
| All | 665 | 100% |

**ICPC-2: International Catalogue of Primary Care version 2**

**Table 4.** GP management actions at the care points. Source: structured case review.

| Management actions | N | Column % |
| --- | --- | --- |
| Drug prescription | 89 | 21% |
| Referral to a specialist | 63 | 15% |
| Advice of over-the-counter drugs | 60 | 14% |
| Renewal of a prescription drug | 36 | 8% |
| Reassurance | 31 | 7% |
| Non-medical advice | 29 | 7% |
| Laboratory testing | 28 | 7% |
| Adjustment of a prescription drug | 20 | 5% |
| Medical imaging | 18 | 4% |
| Physiotherapy | 18 | 4% |
| Other intervention | 14 | 3% |
| Advice to continue over-the-counter drugs | 9 | 2% |
| Other technical examination | 6 | 1% |
| Referral to dentist | 4 | 1% |
| Adjustment of over-the-counter drugs | 1 | 0% |
| All | 426 | 100% |

**Table 5.** Nurse management actions at the care points. Source: structured case review.

| Management actions | N | Column % |
| --- | --- | --- |
| Advice of over-the-counter drugs | 121 | 31% |
| Non-medical advice | 68 | 17% |
| Laboratory testing | 43 | 11% |
| Referral to a specialist | 38 | 10% |
| Reassurance | 31 | 8% |
| Asked the GP to prescribe a drug | 29 | 7% |
| Asked the GP to renew a prescription drug | 20 | 5% |
| Other intervention | 14 | 4% |
| Vaccination | 8 | 2% |
| Advice to continue over-the-counter drugs | 6 | 2% |
| Referral to dentist | 4 | 1% |
| Medical imaging | 3 | 1% |
| Other technical examination | 3 | 1% |
| Adjustment of over-the-counter drugs | 2 | 1% |
| Physiotherapy | 2 | 1% |
| Adjustment of a prescription drug | 1 | 0% |
| All | 393 | 100% |

For 592 out of the 624 nurse consultations (95%), information on the interprofessional workflow was available. The nurses were able to manage half of the cases alone (327, 55%), referred to the GP in 37% (N=217), and consulted a GP (live, by telephone or a dedicated app) for 8% (48).

## 4 Discussion

In this community case report we described how local authorities organized medical services for the group of Ukrainian refugees in a Belgium city through a system of dedicated care points outside of the mainstream health system. The observations provide opportunities to reflect upon the process outcomes coverage and utilization, and content of care, and to reflect upon the lessons both for organization of care for newly arriving migrants and for the mainstream health care system.

These care points combined preventive actions and primary care. Prevention focused on tuberculosis screening and vaccinations, related to the high tuberculosis prevalence in Ukraine. Despite repeated information campaigns a coverage of 30% for prevention is low. Those who made use of the care, had an average of 2,87 consultations per person in 9 months, which seems comparable to the United Nations Refugee Agency indicative statistics of emergency standard health facility utilization of 1-4 consultations per person per year. No information is available about if and where the other 70% of the Ukrainian refugees sought preventive or curative care.

The content of care mostly pertained typical acute primary care illnesses, which is in line with the major population being children and young women and with first phase care in other settings.(19) While Ukrainian refugees in Germany reported high psychological stress and mental health problems, the Antwerp figures don’t suggest that people sought support for this in care points in the first nine months or the healthcare workers did not pick up these problems.(20)

Looking at the registered reasons for encounter and diagnoses, little chronic care was found. This study does not reveal a reason for this finding: is this young refugee population in relatively good health or did the care points not address or detect chronical problems well? Further research is necessary on this subject. The studied contents of care more closely resemble out-of-hours primary care than usual care in primary care practices (primordially minor urgent problems).(21)

Local initiators and city authorities decided upon the organization of care. Higher-level governments supported through financial reimbursement and regulatory flexibility. The vast majority of the work was done by nurses, who were the gatekeeper for GPs consultations. The nurses performed more than half of the consultations autonomously. This community case study shows that it is feasible to set-up a nurse-led care provision supported by GPs for acute common disorders and standardized preventive care for a specific population and their needs. This is also a model used in the regular Belgian system for asylum seekers.(22) These results should be regarded in a broader trend towards integration of nurses in primary care.(23) The studied care points are in line with new organizational care models that take into account interprofessional collaboration and task delegation, in the context of an ageing population and increased pressure on primary care.(24, 25), the care point case study provides a setting with experienced nursed in a more autonomous role then in the current mainstream primary care practices in Belgium. Belgium has got a relative shortage of GPs due to ageing of the GPs and a low attrition of young GPs so installing more nurses in regular primary care might be interesting.(24, 25). The study period was too short and the sample too small to allow for any conclusions regarding the safety of the studied nurse first care model. In addition, the current shortage and lack of suitable training for such cadre of nurses renders it challenging to apply this model to the regular Belgian primary care system at this moment.

Another novelty regarding nurse led care in primary care is the way nurses registered medical diagnoses. Although they slightly differ from the medical diagnoses recorded by GPs, it is compelling that nurses did register medical diagnosis even without involving a GP such as an acute middle ear infection. Nurses did not make use of nursing diagnoses (e.g., acute pain in the ear) which are more problem focused.(26) This finding might be explained by two factors. Firstly, nurses were not specifically trained to make diagnoses. Secondly, the used software was designed for doctors.

While the Ukraine war lingers on and new crises appear, the necessity to think about sustainability becomes urgent. The continuation of the current model rises challenges in terms of comprehensiveness and equity. The current mode is not designed to offer chronic and mental care, while these care needs are expected to increase over time. While the high levels of solidarity of European populations with the Ukraine population contributed to a differentiated approach to Ukraine refugees in the initial phase of the war, this becomes less obvious over time.(27) For instance, other people in vulnerable situations (refugees from other countries, homeless people, illegal substance users, …) do not have equal access to a specialized care point. Local initiatives such as the Antwerp care points can be complementary to other existing structures for refugee services, but coordination and alignment of resources and process is necessary. Currently a long-term follow-up system is necessary as it not feasible to transfer the entire workload to the regular primary care system neither is it desirable to continue the current way of working as it is not adapted to chronic and mental care.

In additional, policy makers need to develop a strategy for integrating migrants into mainstream health service systems in a way the prevents overburdening. Examples from other countries such as Quebec, where the integration process is streamlined through a navigator person for migrants, additional training for GPs and translation services, provide clues for success.(28)

We conclude that local care points were useful for the medical welcome of a sudden influx of refugees. Nurses worked efficiently and only deployed GPs when necessary. Many questions remain concerning the generalizability of these results, safety of the tested care model and continuity of care after the crisis fades away.

## 5 Acknowledgment of conceptual and methodological constraints

This study has the typical limitations of a longitudinal retrospective study without a control group. Comparison of the results to regular Belgian primary care was not possible. Althoug planned, no reliable data concerning the delivery of preventive care was available. The authors were only able to assess the interdiscplinarity follow-up of nurse consultations; no reliable data concerning the interdisciplinary follow-up of physician’s consultations was available.

The patient sample for the retroscpective chart review was randomly drawn but apart from age and sex, the representativeness as compared to the overall population was not studied. Similarly, we do not know whether the population reached by the carepoints was similar to the unreachted population. As reaching a patients was not random, a selection bias is likely. A feasibility sample of 200 patients was chosen, no formal sample size calculation was possible because of the lack of previous studies.

The codebook was jointly created by the authors but afterwards, every case was only judged once because of a lack of funding. The intra- and interrater variability has not been tested. The codebook was based upon variables found in the files, selective registration of certain variables is possible as the patient files were destined for clinical use, not research.

The authors only studied quantitative data. A companiong qualitative study adressing the barriers and facilitators of the studied care model is necessary. Such a study should involve focus groups and in-depth interviews with patients, stakeholders, policy makers and healthcare professionals.

## 7 Author Contributions

All authors participated in the conceptualization and the writing; all authors approved the final version of this manuscript. All authors except KM were involved in the methodology and the investigation. Author SM managed software, managed the project administration, executed the formal analysis, and curated the data. Author JVO validated the formal analysis and wrote the first draft together with SM.

## 9 Acknowledgments

The authors would like to thank all healthcare workers that were willing to collaborate in this research. Google Bard version 2023.10.30 up to version 2023.11.21 from Google AI (https://bard.google.com/chat) was used to rewrite certain paragraphs to increase readability, all content was adapted and approved by the authors.

## 10 Data Availability Statement

The data for this study was collected without individual consent. The raw data (without personal information) is available to other researchers upon reasonable request and after a new request at the Institutional Review Board of the Antwerp University hospital.

